# Field evaluation of a novel, rapid diagnostic assay RLDT, and molecular epidemiology of enterotoxigenic *E. coli* among Zambian children presenting with diarrhea

**DOI:** 10.1101/2022.03.05.22271939

**Authors:** Suwilanji Silwamba, Obvious N. Chilyabanyama, Fraser Liswaniso, Caroline C Chisenga, Roma Chilengi, Gordon Dougan, Geoffrey Kwenda, Subhra Chakraborty, Michelo Simuyandi

**Affiliations:** Centre for Infectious Disease Research in Zambia, Lusaka, Zambia; Department of Biomedical Sciences, School of Health Sciences, University of Zambia, Lusaka, Zambia; Cambridge Institute for Therapeutic Immunology and Infectious Disease, University of Cambridge, UK; Department of International Health, Johns Hopkins University, USA

**Keywords:** LAMP, Enterotoxigenic *E. Coli*, qPCR, Diarrhea, Children, RLDT

## Abstract

**Background:** Enterotoxigenic *Escherichia coli* (ETEC) is one of the top aetiologic agents of diarrhea in children under the age of 5 in low-middle income countries (LMICs). The lack of point of care diagnostic tools for routine ETEC diagnosis results in limited data regarding the actual burden and epidemiology in the endemic areas. We evaluated performance of the novel Rapid LAMP based Diagnostic Test (RLDT) for detection of ETEC in stool as a point of care diagnostic assay in a resource-limited setting.

**Methods:** We conducted a cross-sectional study of 324 randomly selected stool samples from children under 5 presenting with moderate to severe diarrhea (MSD). The samples were collected between November 2012 to September 2013 at selected health facilities in Zambia. The RLDT was evaluated by targeting three ETEC toxin genes [heat labile toxin (LT) and heat stable toxins (STh, and STp)]. Quantitative PCR was used as the “gold standard” to evaluate the diagnostic sensitivity and specificity of RLDT for detection of ETEC. We additionally described the prevalence and seasonality of ETEC.

**Results:** The study included 50.6% of participants that were female. The overall prevalence of ETEC was 19.8% by qPCR and 19.4 % by RLDT. The children between 12 to 59 months had the highest prevalence of 22%. The study determined ETEC toxin distribution was LT (49%), ST (34%) and LT/ST (16%). The sensitivity and specificity of the RLDT compared to qPCR using a Ct 35 as the cutoff, were 90.7% and 97.5% for LT, 85.2% and 99.3% for STh and 100% and 99.7% for STp, respectively.

**Conclusion:** The results of this study suggest that RLDT is sufficiently sensitive and specific and easy to implement in the endemic countries. Being rapid and simple, the RLDT also presents as an attractive tool for point-of-care testing at the health facilities and laboratories in the resource-limited settings.

**Author Summary:** ETEC is one of the top causes of diarrheal diseases in low and middle income countries. The advancement of molecular diagnosis has made it possible to accurately detect ETEC in endemic areas. However, the complexity, infrastructure and cost implication of these tests has made it a challenge to routinely incorporate them in health facilities in endemic settings. The ETEC RLDT is a simple and cost-effective molecular tool that can be used to screen for ETEC in resource limited settings. Here, we described the performance of the RLDT against a qPCR as the gold standard. Our findings showed that the ETEC RLDT performs comparable to the qPCR and would be a suitable screening tool in health facilities in recourse limited settings.

## Introduction

Enterotoxigenic *Escherichia coli* (ETEC) is one of the top ten causes of diarrhea [1] with an estimated 75 million diarrhea episodes annually in children under the age of 5 years. It is also responsible for an estimated 18,700 deaths (9,900–30,659), accounting for ∼ 4.2% (2.2–6.8) of total diarrhea-related deaths [2]. Diarrhea is also associated with long-term consequences of poor growth and cognitive development among children [3,4]. The ETEC disease burden estimates are reportedly lower than the actual cases in endemic areas due to limited diagnostic capacity [5]. In low- and middle-income countries (LMICs), diarrhea remains a wet season disease with enteric pathogens like ETEC playing a fundamental role in warmer and wetter summer months [6,7]. It is therefore crucial to understand the seasonality of ETEC in the region to inform policymakers on prevention and control strategies.

To accurately diagnose ETEC, one needs to first culture stool, isolate *E. coli* colonies and then test if the bacterium produces toxins *(LT, STh, and STp)* through the use of phenotypic assays such as dot blotting or through the use of conventional PCR. Quantitative PCR (qPCR) is performed with purified DNA from stool; although more sensitive; however is technology dependent and is difficult to perform without well-equipped laboratories [8]. The complex nature of the diagnosis leads to (i) long turnaround time, which in turn promotes presumptive treatment that could lead to Antimicrobial Resistance (AMR) [9,10] (ii) increase in cost and labour needed for the detection of ETEC. These are some of the reasons why ETEC is not routinely tested in resource-limited settings.

The complexity of these diagnostic methods results in the underestimation of the burden of ETEC because countries where the infection is endemic cannot afford the infrastructure and expertise required for this [11]. To develop an effective program for control and prevention, accurate burden data is important [12]. In resource-limited settings, there is a need for a simple, readily available method that can be used to detect ETEC in minimally equipped laboratories and health settings.

*Chakraborty et al* previously developed a simple diagnostic assay, a Rapid LAMP based Diagnostic Test (RLDT) for detection of ETEC, which is based on Loop mediated Isothermal Amplification (LAMP) [13,14]. The RLDT detects ETEC directly from stool in <1 hour. In this study, we field evaluated the RLDT in Zambia compared with qPCR, using previously collected stool samples. We also described the prevalence of ETEC infections among the Zambian children presenting with moderate to severe diarrhea (MSD) as well as asymptomatic cases, at the outpatient clinics.

## Materials and Methods

### Study design

This was a retrospective study using 324 randomly selected samples from 1500 stored stool samples collected at various health facilities in the Lusaka district of Zambia. These samples were collected between November 2012 to September 2013 from a previous rotavirus vaccine effectiveness study [15] as shown in Figure 1. clinical information including diarrhea severity, social demographic data were collected from study participants.

**Figure 1:**
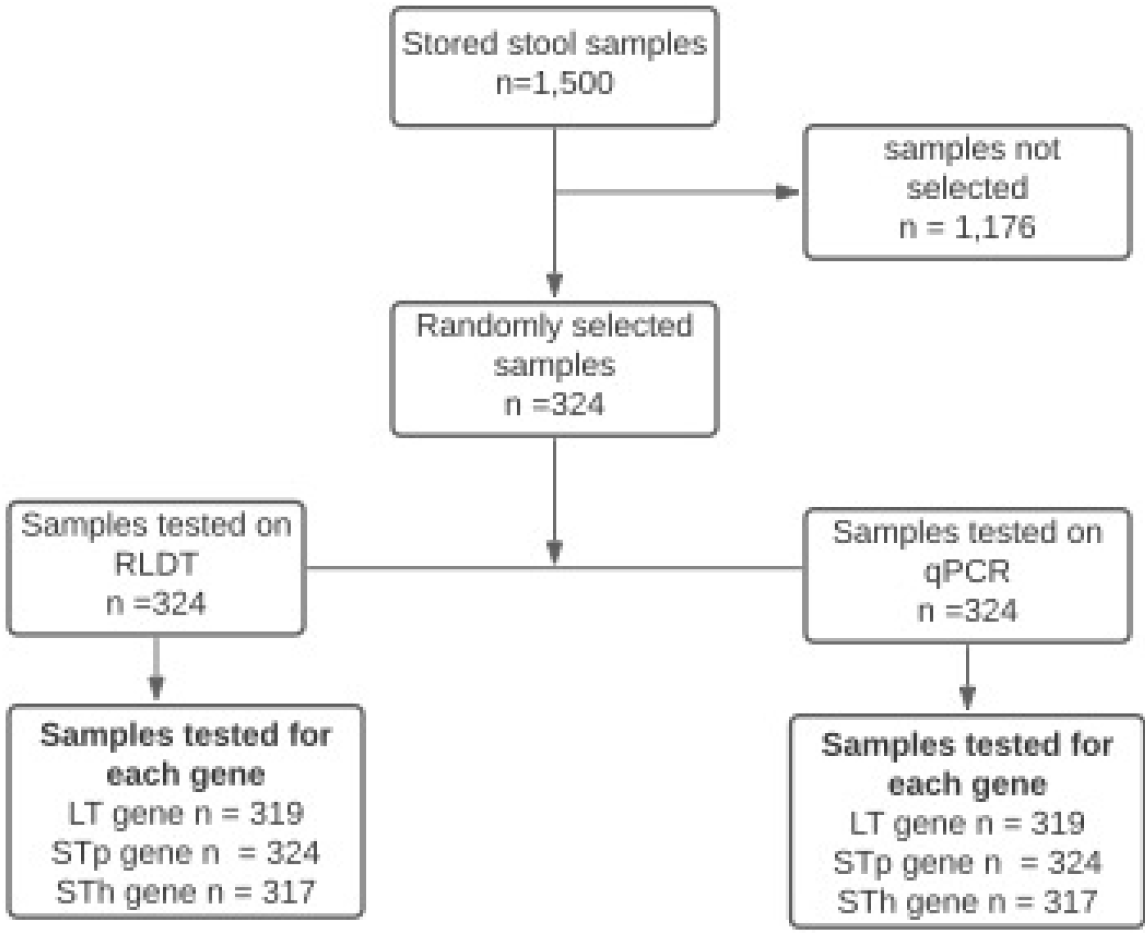
Study participant flow chart.

#### Randomization of stool samples before selection

An independent statistician was tasked to randomly select 324 retrospectively collected samples. This set of samples represented stool samples with equal distribution of sex and under 5 age groups. The statistician also stratified the participant samples with a 2:1 ratio of symptomatic and asymptomatic diarrhea cases.

### Laboratory Assays

#### Sample Processing Collection and Storage

Samples with collected clinical information of moderate to severe diarrhea and asymptomatic representation were sorted, separated and stored at -80°C before testing.

#### ETEC –RLDT Training

A team from the Johns Hopkins University, United States, Baltimore travelled to Zambia to train staff at the Centre for Infectious Diseases Research in Zambia (CIDRZ) on RLDT and quantitative PCR (qPCR) assays in an effort to build laboratory capacity. The RLDT and qPCR training duration lasted 2 weeks and was followed up by evaluation of performance of the staff. The staff at CIDRZ, found that the RLDT assay is simple to use and the assay could be performed by non-skilled laboratory personnel.

#### ETEC –RLDT Assay

RLDT assays were conducted directly from the frozen stool samples using the RLDT kit as described by Chakraborty et al [13]. In short, samples were added to a sample processing tube with lysis buffer followed by heat lysis. The processed samples were then added to the ETEC RLDT lyophilized reaction tube (LRT) strips. Each strip consisted of 8 tubes, organized as two reaction tubes each for LT, STh and STp genes. One reaction tube was added as the RLDT inhibitor control [14]. The strips were run for 40 minutes in a real time fluorometer reader (Agdia Inc, IN, USA). The results were read as positive/negative by the reader.

#### qPCR Assay

##### Nucleic Acid extraction

About 100-150mg of bulk stool were added to SK38 bead tubes (Bertin Technologies, Montigny, France) containing lysis buffer (bioMérieux, Marcy I’Etoile, France). The stool suspension was vortexed for 5 minutes, allowed to stand at room temperature for 10 to 15 minutes, then centrifuged at 14,000 rpm for 2 minutes to pellet stool material. About 200μl of the supernatant were transferred into a nuclease-free 1.5ml microcentrifuge tube for extracting nucleic acid using a QIAamp DNA Mini Kit (Qiagen, Hilden, Germany) according to the manufacturer’s instructions.

##### qPCR Amplification

The 25μl reaction mixtures contained 12.5ul Quantitech SYBR Green Master mix (Qiagen, Hilden, Germany), 1uM primer mix 5ul, PCR grade water 5ul (Invitrogen, USA) and 2.5ul of samples. PCR was carried out for 40 cycles of 95°C for 15s and 60°C for 1 min [16]. qPCR cycling conditions were run on the RotogeneQ platform (Qiagen, Hilden Germany). Cut-off for the determination of ETEC positives was set as Ct35 as was done in previous studies [5]. Each sample was run at a minimum in duplicate, and results were averaged.

Chakraborty et al previously has established the limit of detection (LOD) of RLDT for ETEC genes LT, STh and STp using stool samples spiked with reference ETEC strain (PNTD-D-21-01199R1, PNTD-D-21-01198R1). The LOD was 9×10^4^ CFU/g of stool which corresponds to qPCR Ct of 28.2, 28.6 and 30.07 for LT, STh and STp respectively). Therefore, we also evaluated the performance of the RLDT using this LOD (Ct 28) as the cut off. (S2 Table).

## Definitions

Diarrhea (Symptomatic) was defined as the primary caregiver reporting that the child had three or more loose stools within 24 hours.

An asymptomatic case was defined as a child presenting to a health facility with other non-diarrhea complications.

## Statistical analysis

A minimum sample size of 324 with an assumed ETEC prevalence of 40.7% [18] produces a two-sided 95% sensitivity confidence interval with a width of 12% when the sample sensitivity is at least 85% and the two-sided 95% specificity confidence interval with a width of 5% when sample specificity is at least 0.95%. We calculated summary statistics for all baseline variables. Proportions and median (IQR) were used to express categorical and continuous variables. A Chi-square test was used to determine the association between ETEC positivity and baseline characteristics.

Statistical analysis significance was set at p-value <0.05 and data were analysed using Stata version 16.0 (StataCorp LLC, College Station, Texas). The correlation of ETEC monthly positivity frequencies was assessed to determine seasonality.

A sample was considered positive for ETEC, when at least one of the ETEC genes, LT, STh or STp was positive. To compare RLDT with qPCR, a Ct value cut off of 35 was used. Any sample with Ct value of 35 or less by qPCR was considered as true positive. To avoid incorrectly determining some samples to be false positive by RLDT, samples with Ct-values greater than 35 detectable by both qPCR and RLDT were also included as true positive. We also compared RLDT with qPCR with the Ct value cut off of 28.

## Ethics

For this study, Ethics and regulatory approvals were sought from the University of Zambia Biomedical Research Ethics Committee (UNZABREC) reference number 009-10-18 and the National Health Regulatory Authority (NHRA). Samples were de-identified and given a unique study participant number to maintain confidentiality.

## Results

### Study design is described in a flow chart (Fig 1)

#### Social demographics and prevalence

A total of 324 samples with a mean age of about 30months were included in the analysis, 50.9% were female, 28.4% were asymptomatic, with 3.1% of the symptomatic cases presenting with severe disease according to a modified versikari severity scoring (Denise et al unpublished) [19]. Overall, ETEC prevalence was about 19% with both the assays, RLDT and qPCR and the highest prevalence was observed in children between 12-59 months of about 22% (Table 1.)

**Table 1.** Baseline characteristics by qPCR/ RLDT positivity.

|  | Total samples tested n (%) | Positive by RLDT n (%) | p value | Positive by qPCR n (%) | p value |
| --- | --- | --- | --- | --- | --- |
| **Age** |  |  |  |  |  |
| <12 months | 159 (49.1) | 26 (16.4) | 0.41 | 26 (16.4) | 0.44 |
| 12-23 months | 37 (11.4) | 9 (24.3) |  | 8 (21.6) |  |
| 24-59 months | 98 (30.2) | 21 (21.4) |  | 22 (22.4) |  |
| missing * | 30 (9.3) | 7 (23.3) |  | 8 (26.7) |  |
| **Sex** |  | 63 |  | 19.75 |  |
| Male | 152 (46.9) | 34 (22.4) | 0.29 | 34 (22.4) | 0.35 |
| Female | 165 (50.9) | 29 (17.6) |  | 30 (18.2) |  |
| missing * | 7 (2.2) | 0 (0) |  | 0 (0) |  |
| **Symptomatic** |  |  |  |  |  |
| No | 92(28.4) | 12(13) | 0.07 | 13(14.1) | 0.092 |
| Yes | 227(70.1) | 51(22.5) |  | 51(22.5) |  |
| missing * | 5(1.5) | 0(0) |  | 0(0) |  |
| **Severity** |  |  |  |  |  |
| Mild /Moderate | 287 (88.6) | 50 (17.4) | 1.0 <sup>1</sup> | 51 (17.8) | 0.70 <sup>1</sup> |
| Severe | 10 (3.1) | 1 (10) |  | 2 (20) |  |
| missing * | 27 (8.3) | 12 (44.4) |  | 11 (40.7) |  |
| **Wash** |  |  |  |  |  |
| Adequate | 213 (65.7) | 40 (18.8) | 0.15 | 40 (18.8) | 0.70 |
| Inadequate | 62 (19.1) | 11 (17.7) |  | 13 (21) |  |
| missing | 49 (15.1) | 12 (24.5) |  | 11 (22.4) |  |
| **Total** | **324 (100)** | **63 (19.4)** |  | **64 (19.8)** |  |

NOTE: Chi square test was used to compare the association of baseline characteristics such as age, sex, Note: diarrhea severity and wash data against RLDT and qPCR ETEC positivity. P values less than 0.05 showing a statistically significant difference. \*Statistical significance (P < 0.05).

#### Performance of the RLDT against qPCR

The performance of the RLDT against qPCR is shown in Table 2. The prevalence of ETEC was 19.8% by qPCR and 19.4% by RLDT. The evaluation of the LT, STh and STp toxin genes sensitivity and specificity of the RLDT using a Ct 35 value cut-off with a 95% confidence interval were, 90.7% (77.9-97.4) and 97.5% (94.8-99); (85.2% (66.3-95.8) and 99.3% (97.5-99.9); and 100% (59.0-100) and 99.7% (98.3-100)) respectively. With the Ct cut off of 28, the sensitivity and specificity were higher (Table S1)

**Table 2.** Performance of RLDT against qPCR.

| Ct≤35** | Number of samples tested | Samples positive by RLDT | Samples positive by qPCR | False positive | False negative | Sensitivity (95% CI) | Specificity(95%CI) |
| --- | --- | --- | --- | --- | --- | --- | --- |
| LT | 319 | 46 | 43 | 7 | 4 | 90.7 (77.9 -97.4) | 97.5 (94.8 - 99.0) |
| STp | 324 | 8 | 7 | 1 | 0 | 100 (59.0 -100.0) | 99.7 (98.3-100.0) |
| STh | 317 | 25 | 27 | 2 | 4 | 85.2 (66.3-95.8) | 99.3 (97.5-99.9) |

Note: 35\*\* a Ct value cutoff for both qPCR and the ETEC RLDT, CI = Confidence Interval

#### Performance of RLDT against qPCR by the clinical representation and AUC analysis

The performance of the RLDT against qPCR by the participants’ clinical representation is shown in Table 3. The evaluation of symptomatic participants of the *LT, STh and STp* toxin genes sensitivity and specificity of the RLDT using the CT value cutoff of 35 with a 95% confidence interval was 91.4% (76.9-99.7), and 96.8% (93.2-98.8), 85.7% (63.7-97.0) and 99% (96.5-99.9) and 100% (54.1-100) and 100% (98.3-100) respectively. Similar results observed when asymptomatic cases were evaluated for sensitivity and specificity of *LT, STh and STp* (87.5% (47.4-99.7) and 98.8% (93.5-100), (83.3% (35.9-99.6) and 100% (95.8-100)) and (100% (2.5-100) and 98.9% (94-100) respectively. A comparison of the ETEC RLDT to qPCR tests for each target gene using Area Under the Curve (AUC) analysis to evaluate the performance of the two instruments. From the analysis, we found there was no significant difference between the ETEC RLDT to qPCR (Figure 2)

**Table 3.** Performance of RLDT against qPCR by the clinical state of participants.

| Ct≤35** | Clinical Status | Number of samples tested | Samples positive by RLDT | Samples positive by qPCR | False positive | False negative | Sensitivity(95%CI) | Specificity(95%CI) |
| --- | --- | --- | --- | --- | --- | --- | --- | --- |
| LT |  |  |  |  |  |  |  |  |
|  | Asymptomatic | 92 | 8 | 8 | 1 | 1 | 87.5(47.4-99.7) | 98.8(93.5-100) |
|  | Symptomatic | 224 | 38 | 35 | 6 | 3 | 91.4(76.9-98.2) | 96.8(93.2-98.8) |
| STp |  |  |  |  |  |  |  |  |
|  | Asymptomatic | 92 | 2 | 1 | 1 | 0 | 100.0(2.5-100.0) | 98.9(94.0-100.0) |
|  | Symptomatic | 227 | 6 | 6 | 0 | 0 | 100 (54.1-100.0) | 100.0(98.3-100.0) |
| STh |  |  |  |  |  |  |  |  |
|  | Asymptomatic | 91 | 5 | 6 | 0 | 1 | 83.3(35.9-99.6) | 100(95.8-100.0) |
|  | Symptomatic | 223 | 20 | 21 | 2 | 3 | 85.7(63.7-97.0) | 99.0 (96.5-99.9) |

Note: 35\*\* a Ct value cutoff for both qPCR and the ETEC RLDT, CI = Confidence Interval

**Fig 2.**
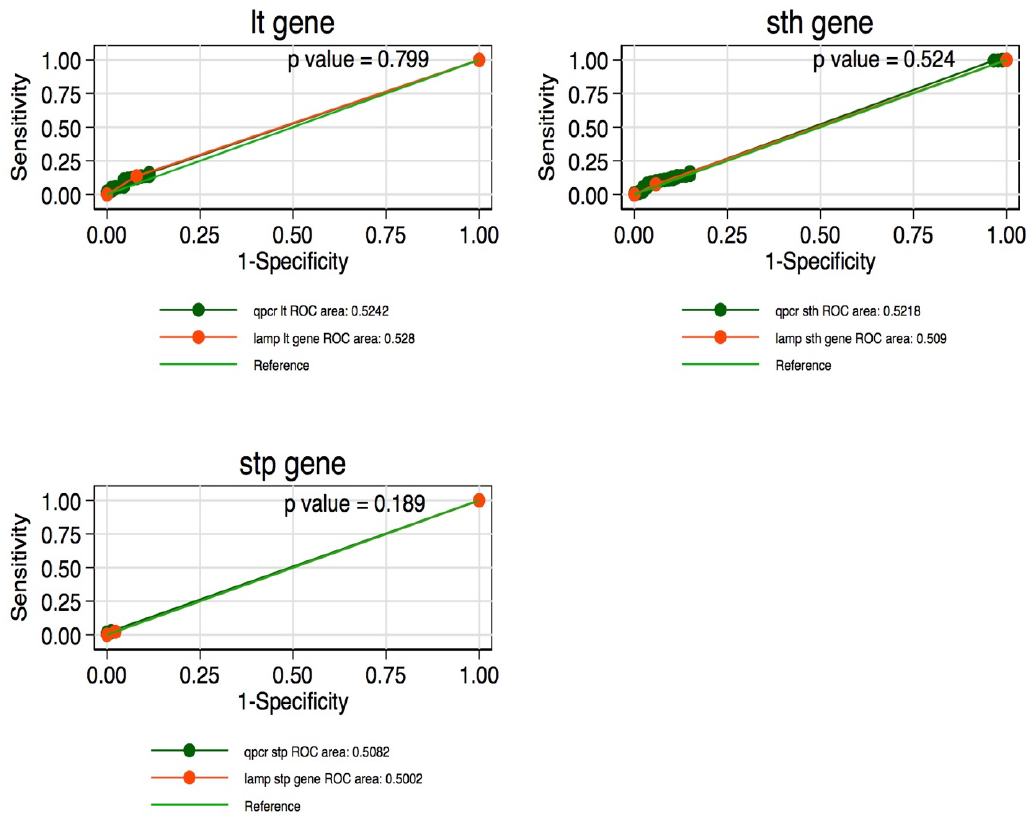
Comparison of RLDT to qPCR tests for each gene using AUC analysis. Note: *Statistical significance (p < 0.05)

#### ETEC toxin gene distribution

ETEC expressing only the heat Labile toxin (LT) had a frequency of 49% being the dominant expressed gene. Followed by 34% of strains expressing only the Heat stable toxin (ST) genes. The frequency of ETEC expressing the combination of both LT/ST toxins was 16% as shown in figure 3

**Fig 3.**
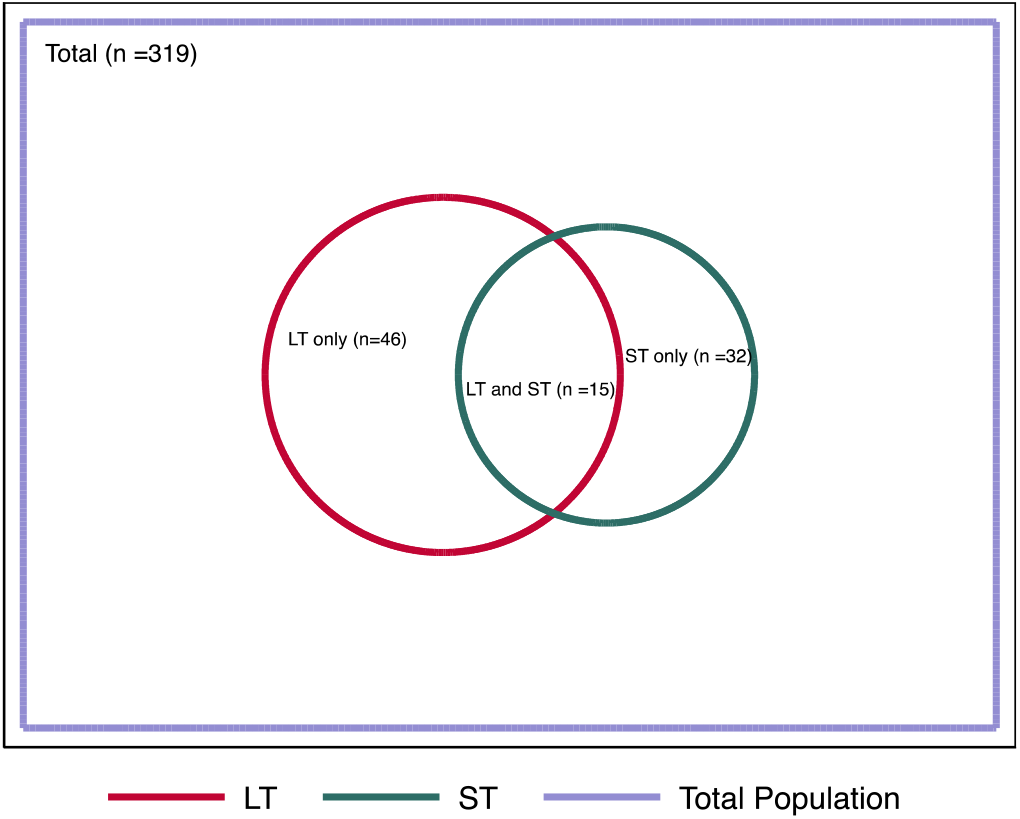
Shows the distribution of ETEC toxins.

#### Seasonality

We observed a seasonal trend of ETEC over 12 months with high positivity rates between December to February (warm, rainy season) and a minor peak between April and May (dry season) (Figure S1) S1 figure.

## Discussion

This study is the first field evaluation of ETEC RLDT and establishes that it performed equally as the qPCR, as demonstrated by the specificity, sensitivity and AUC curves for each toxin gene *LT, STh* and *STp*. The performance of the RLDT was similar among ETEC positive diarrhea and asymptomatic cases. These findings are important as they support the use of the RLDT for screening for ETEC among children presenting with diarrhea at health facilities. In addition, its turnaround time and simplicity (not requiring skilled laboratory personnel for testing and results interpretation) makes it ideal for resource-limited settings. The RLDT could also be implemented in these countries for ETEC disease surveillance which is crucial to obtain meaningful disease burden data to inform policymakers and healthcare professionals for developing control and prevention programs. Similar studies which aimed at assessing LAMP platforms sensitivity and specificity against qPCR for the detection of *Mycoplasma pneumonia* [20] and *Leptospira spp* [21] found that both the LAMP assays had good sensitivity and specificity (99.1% and 100.0%) and (96.8% and 97.0%), respectively. These studies also concluded that the LAMP platforms are easy to use and comparable to the qPCR, as shown in this study.

We determined that across the stratified age groups, the children between 12 to 59 months were at the highest risk of getting ETEC infection with prevalence of ∼ 22%. The overall prevalence of ETEC under 5 years old, in this study was ∼19%. The isolation rates of ETEC in our study is similar to previous studies that have reported the prevalence of ETEC in developing countries from Bangladesh, Turkey, Peru, Mexico, Egypt, Argentina, India, Nicaragua, and Tunisia which indicated a rate of 18-38% in children [22–26]. However, the ETEC prevalence in our study was lower than what was reported in a previous study (40.7%) conducted in Zambia [18] using Luminex Magpix GPP panel which uses x-TAG technology. This could be attributed to the different in testing platforms, technology and sensitivity of the assays.

The seasonal prevalence observed in our study is similar to what was reported in Kenya [27] which reported the seasonal variation of enteric bacterial pathogens among the hospitalized children with diarrhea. ETEC infections were found all year round with an increase during the warm rainy season and dry seasons. [28]. This information is critical to inform policymakers and healthcare professionals to develop control and prevention programs including when to deploy the ETEC vaccines.

We also found that in Lusaka, Zambia, among the circulating ETEC strains, the LT-ETEC strains was the highest followed by ST-ETEC and LT+ST-ETEC strains. Only 6% of the ETEC strains were STp-ETEC. A similar distribution of toxin genes among ETEC strains was reported from Bolivia (LT 70%, LT+STh 23% and STh 7%) [29]. Michelo et al, also reported similar results, LT+STh being the most common toxin combination and LT+STh+STp being the least common in Zambia[30]. This suggests that vaccines such as ETVAX could be effective for this population and region.

### Strengths

This study has several strengths firstly the results of this study were obtained from samples that were collected from several health facilities across the city of Lusaka, which means that our findings can be generalized across Lusaka Province. In addition, this study included symptomatic and asymptomatic cases that were stratified by age. This sampling method accounted for some biases in the computation of the true prevalence. We conducted qPCR assays with samples run in duplicate using average CT values increasing the accuracy of the study results. This study’s novelty evaluates the ETEC RLDT platform, demonstrating a successful set-up in a resource-limited setting, which is comparable to qPCR.

### Limitations

This study has a couple of limitations which include but are not limited to (i) The cross-sectional design only provides prevalence data but does not provide incidence data, which is critical for vaccine trial design and planning control and prevention efforts. The two assays qPCR and RLDT, compared here, used different sample preparation methods. RLDT was done directly from the stool with minimum sample treatment; qPCR was done from purified DNA; Therefore, the sensitivity of these assays depends not only on the amplification technology but also on the starting material. In addition, while qPCR is quantitative, RLDT is semi-quantitative.

## Conclusion

We found that the RLDT performed comparable to the qPCR assay. Additionally, the observed specificity and sensitivity are high enough to suggest that the RLDT could be used in a field setting to rapidly detect ETEC among patients presenting with diarrhea in the health facilities. This study justifies a broader application of the RLDT as a simple and rapid diagnostic test for ETEC in the endemic countries where such simple assays are critically needed. We also determined that LT-ETEC and ST-ETTEC strains were highly prevalent and ETEC positivity was highest in the warm rainy season.

## Data Availability

The underlying data set cannot be made publicly available because it contains human research participant data; however, it can be made available to any interested researchers upon request. The Centre for Infectious Disease Research in Zambia (CIDRZ) Ethics and Compliance Committee is responsible for approving such requests. To request data access, one must write to the Secretary to the Committee mentioning the intended use for the data, contact information, a research project title, and a description of the analysis being proposed as well as the format it is expected. The requested data should only be used for purposes related to the original research or study. The CIDRZ Ethics and Compliance Committee will normally review all data requests within 48 to 72 hours (Monday to Friday), and provide notification if access has been granted or additional project information is needed.

## Author Contributions

SS and MS drafted the original manuscript, MS worked on the Conceptualization and design of the study, SS and FL performed laboratory procedures and generated the laboratory information. OC and SC performed the statistical analysis for the study. SS, RC, SC, GD, GK, CCC and MS thoroughly reviewed and edited the article. Funding of the study MS SC GD. All the co-authors reviewed and agreed on the final submission.

## Acknowledgements

We thank all clinical, data, and laboratory staff at the Centre for Infectious Disease Research in Zambia for their contributions to this work. We also acknowledge the training and material contributions from the Johns Hopkins University in Baltimore, USA.

## Data sharing statement

The underlying data set cannot be made publicly available because it contains human research participant data; however, it can be made available to any interested researchers upon request. The Centre for Infectious Disease Research in Zambia (CIDRZ) Ethics and Compliance Committee is responsible for approving such requests. To request data access, one must write to the Secretary to the Committee (via.) mentioning the intended use for the data, contact information, a research project title, and a description of the analysis being proposed as well as the format it is expected. The requested data should only be used for purposes related to the original research or study. The CIDRZ Ethics and Compliance Committee will normally review all data requests within 48–72 hours (Monday-Friday), and provide notification if access has been granted or additional project information is needed.

